# Resource-Aware Conditional Diffusion for CT-to-PET Translation Supporting Rural Oncology Imaging

**DOI:** 10.64898/2026.03.09.26347907

**Authors:** Srijita Khatua

**Affiliations:** IIT (ISM) Dhanbad

**Keywords:** CT-to-PET Translation, Diffusion Models, Rural Healthcare, Oncology Screening, SUV Preservation

## Abstract

Access to positron emission tomography (PET) remains limited in rural and low-resource healthcare settings due to high infrastructure cost and radiotracer logistics. This restricts early oncologic screening in underserved populations. The study proposes a rural-optimized conditional diffusion framework for synthetic PET generation directly from widely available CT scans. The architecture employs a two-stage residual design consisting of a lightweight coarse predictor followed by computationally efficient diffusion refinement with reduced timesteps and deterministic sampling. A multi-objective SUV-aware loss ensures metabolic consistency. To emulate rural deployment conditions, this study simulates low-dose noise, Hounsfield unit miscalibration, and resolution degradation. Clinical validation demonstrates strong structural fidelity (SSIM 0.81) and stable SUVmean preservation. Domain-matched training achieves SUVmax error as low as 0.61. Cross-dataset analysis highlights the importance of SUV harmonization for robust rural deployment. This work presents a resource-sensitive AI framework supporting equitable oncology screening in rural healthcare systems.

**Highlights:**

- Two-stage residual conditional diffusion for CT-to-PET translation.
- SUV-aware multi-objective optimization preserves metabolic biomarkers.
- Few-shot adaptation improves cross-dataset SUV calibration.

## 1. Introduction

Cancer remains a major public health challenge in India and other low- and middle-income countries, where rising incidence and mortality are compounded by healthcare disparities. This challenge accounts for a significant proportion of cancer-related deaths, with socioeconomic status, environmental exposure, and access to diagnostic imaging strongly influencing outcomes (Gunani et al., 2025). Advanced imaging modalities such as positron emission tomography (PET) play a central role in cancer staging, response evaluation, and treatment planning. However, equitable access to PET imaging remains limited. Nowadays, CT-PET has evolved into a cornerstone modality for oncologic care, enabling detection of metabolic dysregulation prior to anatomical changes (Khan, 2016). Despite its clinical utility, the expansion of PET infrastructure has been constrained by high scanner costs, radiotracer logistics, and workforce limitations.

The consequences of limited PET access are clinically significant. Patients from lower socioeconomic backgrounds are less likely to undergo advanced imaging such as CT or PET due to financial constraints, often leading to delayed diagnosis and suboptimal treatment outcomes (Gunani et al., 2025). In addition, disparities in imaging awareness and understanding remain evident in rural populations, with knowledge gaps particularly pronounced for advanced modalities such as PET (Jamwal and Kaushal, 2025). Together, these infrastructural and educational barriers hinder timely metabolic assessment and early cancer detection in underserved communities.

While strengthening PET infrastructure remains a priority (Khan, 2016; Kachroo et al., 2025), alternative computational strategies may complement long-term planning efforts. With CT scanners being more widely available across district hospitals and semi-urban centers, synthetic PET generation from CT presents a potential pathway toward scalable metabolic screening support. Such approaches may not replace true PET imaging but could assist in preliminary triage, metabolic trend estimation, and decision support in resource-constrained settings.

Despite progress in PET image generation and cross-modality translation, several limitations persist:

- Computationally intensive generative frameworks not optimized for deployment in resource-constrained settings.
- Limited incorporation of oncology-specific quantitative constraints such as SUV consistency.
- Insufficient evaluation under domain shift or variable acquisition conditions.
- Minimal focus on equitable deployment in rural and underserved populations.

To address these gaps, the study proposes a rural-optimized conditional diffusion framework for CT-to-PET translation. Unlike prior GAN-based or generic diffusion models, our approach integrates reduced diffusion steps, deterministic sampling, residual learning stabilization, and SUV-aware multi-objective optimization to promote metabolic fidelity while maintaining computational feasibility.

## 2. Related Work

### 2.1. Artificial Intelligence in PET Imaging

Artificial intelligence (AI) has increasingly been applied to positron emission tomography (PET) across diagnostic and image generation tasks. A comprehensive review by Matsubara et al. (Matsubara et al., 2022) categorizes deep learning applications in PET into three major themes: (1) denoising and recovery of full PET data from noisy acquisitions, (2) PET reconstruction and attenuation correction, and (3) PET image translation and synthesis. These approaches primarily employ convolutional neural networks (CNNs) and generative adversarial networks (GANs) to enhance image quality, reduce acquisition time, or enable cross-modality translation. While these studies demonstrate the feasibility of deep learning for PET image generation, most focus on improving image realism or reconstruction accuracy rather than enforcing oncology-specific quantitative constraints such as standardized uptake value (SUV) consistency. Given that PET provides quantitative metabolic information critical for cancer staging and response evaluation, preserving SUV fidelity remains an important challenge.

### 2.2. PET Radiotracer Translation and Clinical Standardization

Beyond image synthesis, translational standardization in PET imaging remains a key concern for oncology. McAteer et al. (Mcateer et al., 2025) highlight the complexities associated with clinical translation of PET radiotracers, emphasizing the need for standardized methodologies, reproducibility, and streamlined regulatory frameworks. Their consensus-based recommendations underscore the importance of harmonization, reproducibility assessment, and stakeholder engagement to accelerate clinical implementation. These translational considerations are particularly relevant when introducing AI-based PET generation frameworks. Any synthetic PET methodology must be evaluated not only for structural similarity but also for quantitative reliability, reproducibility, and potential integration into existing oncology workflows.

### 2.3. CT-to-PET Image Translation

Image-to-image translation frameworks have been increasingly investigated for generating PET images from CT scans. GAN-based approaches have demonstrated strong structural similarity but often struggle with metabolic intensity calibration. Guarrasi et al. (Guarrasi et al., 2025) proposed a district-specific GAN framework for whole-body CT-to-PET translation, segmenting anatomical regions (head, trunk, arms, and legs) and training region-specific GANs to address anatomical heterogeneity. Their method achieved improved quantitative performance compared to non-segmented GAN baselines, particularly in paired Pix2Pix settings. However, the primary objective remained structural fidelity and image realism rather than enforcing quantitative metabolic constraints. More recently, Nguyen et al. (Nguyen et al., 2025) introduced CPDM, a domain-knowledge-guided conditional diffusion model for CT-to-PET translation. CPDM employs a Brownian Bridge diffusion process to directly map CT to PET distributions and incorporates two domain-informed conditional maps: an Attention map to guide focus on clinically relevant regions, and an Attenuation map to improve PET correction and diagnostic consistency. The authors also presented one of the largest paired CT-PET datasets to date and demonstrated performance improvements over GAN-based and diffusion-based baselines across multiple metrics.

These works establish diffusion modeling as a promising direction for CT-to-PET translation. However, existing frameworks primarily emphasize large-scale training or domain-guided conditioning mechanisms, with limited focus on deployment feasibility under resource-constrained settings or evaluation under rural healthcare variability.

## 3. Methodology

The study proposes a rural-optimized two-stage conditional diffusion framework for CT-to-PET translation. Given a CT image *X* ∈ ℝ^*H×W*^, the objective is to generate a synthetic PET image *Ŷ* that preserves both structural similarity and metabolic fidelity. The overall formulation is:

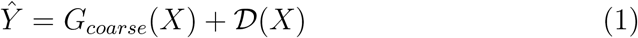

where *G*_*coarse*_ is a lightweight convolutional network and 𝒟 is a conditional diffusion-based residual refinement model. This residual formulation stabilizes training and reduces diffusion burden.

### 3.1. Stage 1: Coarse PET Prediction

A lightweight convolutional network generates an initial PET estimate:

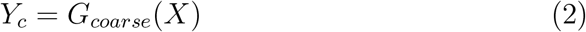

The coarse predictor is implemented as a shallow three-layer convolutional network designed for computational efficiency. Specifically, it consists of two intermediate convolutional layers with 64 feature channels each, followed by a final projection layer producing a single-channel PET estimate. All convolutional layers use 3 × 3 kernels with padding to preserve spatial resolution. Rectified Linear Unit (ReLU) activations are applied after the first two layers, while the final layer outputs a linear intensity prediction.

Functionally, the coarse network captures global anatomical-to-metabolic correspondence between CT intensity patterns and PET uptake distributions. While it does not model fine-grained metabolic heterogeneity, it provides a stable structural baseline that reduces the burden on the subsequent diffusion refinement stage. By predicting a coarse approximation of PET activity, the diffusion module is tasked only with modeling the residual metabolic details rather than the full CT-to-PET mapping.

### 3.2. Stage 2: Conditional Diffusion Refinement

This work employs a conditional Gaussian diffusion process with reduced timesteps *T* = 200. Here Forward diffusion process:

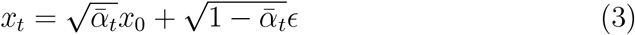

where *x*_0_ = *Y* −*Y*_*c*_ (residual target), *ϵ*∼ *𝒩* (0, 1),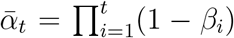 model predicts noise conditioned on CT:

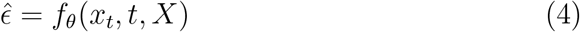

Here training objective is

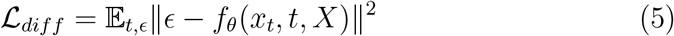

### 3.3. Stage 2: Conditional Diffusion Refinement

The study employs a conditional Gaussian diffusion process with reduced timesteps *T* = 200, significantly lower than conventional diffusion models (typically *T* = 1000), to reduce computational burden and enable faster inference suitable for low-resource deployment. Rather than modeling the full PET distribution directly, this work applies diffusion to the residual between the ground-truth PET image *Y* and the coarse prediction *Y*_*c*_:

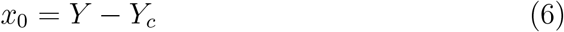

This residual formulation simplifies the generative task by allowing the diffusion model to focus on fine-grained metabolic corrections rather than global anatomical structure. Here the forward diffusion process gradually perturbs the residual with Gaussian noise:

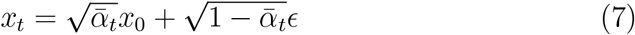

where *ϵ* ∼ *𝒩* (0, 1) is Gaussian noise, *β*_*t*_ is a linear noise schedule, *α*_*t*_ = 1 − *β*_*t*_, 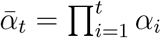. As *t* increases, the residual becomes progressively noisier until it approaches an isotropic Gaussian distribution. The conditional diffusion model predicts the noise component conditioned on the CT image:

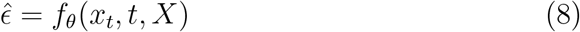

where *f*_*θ*_ is implemented as a lightweight conditional UNet. The training objective minimizes the mean squared error between true and predicted noise:

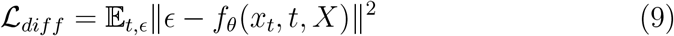

During inference, this work adopts a deterministic DDIM(Denoising Diffusion Implicit Models) sampling update rule with step size 2, effectively reducing the number of reverse diffusion steps from 200 to 100. This significantly lowers inference time while preserving image fidelity. To improve training stability and inference robustness, this work maintain an exponential moving average (EMA) of model parameters. Let *θ* denote the current model parameters and *θ*_*ema*_ denote the averaged parameters. After each update, EMA parameters are updated as:

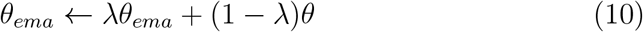

where *λ*∈ [0, 1) is the decay factor set to 0.999.

At inference time, this work uses *θ*_*ema*_ rather than the instantaneous parameters *θ*, resulting in improved convergence smoothness and reduced artifact generation. This stabilization strategy is particularly beneficial in low-resource training settings, where smaller batch sizes and limited computational capacity may otherwise increase optimization noise.

### 3.4. SUV-Aware Multi-Objective Optimization

Positron emission tomography (PET) provides quantitative measurement of radiotracer uptake, commonly expressed using the standardized uptake value (SUV). SUV is a semi-quantitative metric defined as the ratio between tissue radioactivity concentration and the injected tracer dose normalized by body weight. It reflects glucose metabolism in oncologic imaging and plays a central role in tumor staging, therapy response assessment, and prognostic evaluation (Adams et al., 2010).

However, SUV measurements are subject to biological and technical variability. As discussed by Brendle et al. (Brendle et al., 2015), SUV can be influenced by patient-specific factors such as body weight, blood glucose level, uptake time, and respiratory motion, as well as technical parameters including scanner calibration, reconstruction algorithms, matrix size, smoothing filters, and the use of point-spread function (PSF) or time-of-flight (TOF) reconstruction. Variations in region-of-interest (ROI) definition and thresholding strategies can further induce SUV differences of up to 50%. Therefore, while SUV is clinically indispensable, its reliability depends strongly on standardization and consistent quantification methodology. Different SUV measurements are commonly used in oncology:

- **SUVmax**: the maximum voxel intensity within a lesion, representing the highest metabolic activity. It is highly sensitive to image noise and reconstruction parameters.
- **SUVmean**: the average SUV within a delineated region of interest (ROI), dependent on lesion segmentation and subject to inter-observer variability.
- **SUVpeak**: the average SUV within a fixed small region around the hottest voxel, designed to reduce sensitivity to noise compared to SUVmax.

Given the clinical importance of SUV metrics, optimizing only pixel-wise similarity may result in structurally realistic but metabolically inconsistent synthetic PET images. To enforce oncology-relevant fidelity, the study incorporates SUV-aware constraints into the training objective.

The total loss is defined as:

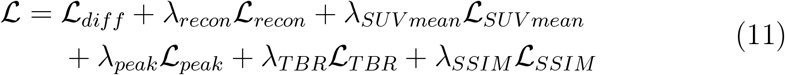

Where

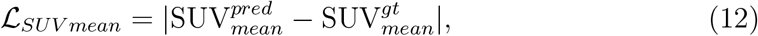

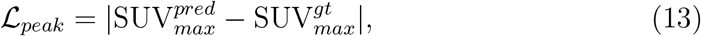

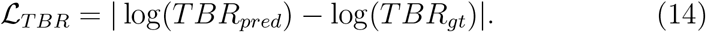

The *ℒ*_*SUV mean*_ term enforces global metabolic consistency across the scan, reducing bias in overall tracer distribution. The peak loss *ℒ*_*peak*_ encourages accurate preservation of high-uptake tumor regions, which are critical for staging and response monitoring.

This study also includes a tumor-to-background ratio (TBR) constraint, defined as the ratio of tumor SUV to background SUV (Rogasch et al., 2015). The logarithmic formulation stabilizes optimization and reduces sensitivity to extreme ratios. TBR alignment is clinically relevant because therapy response evaluation often depends on the relative contrast between the tumor and the surrounding tissue rather than the absolute intensity alone. By explicitly incorporating SUV-aware losses, the model promotes metabolic fidelity beyond structural similarity, aligning synthetic PET generation with oncology-specific quantitative requirements.

#### Algorithm 1 Training Algorithm

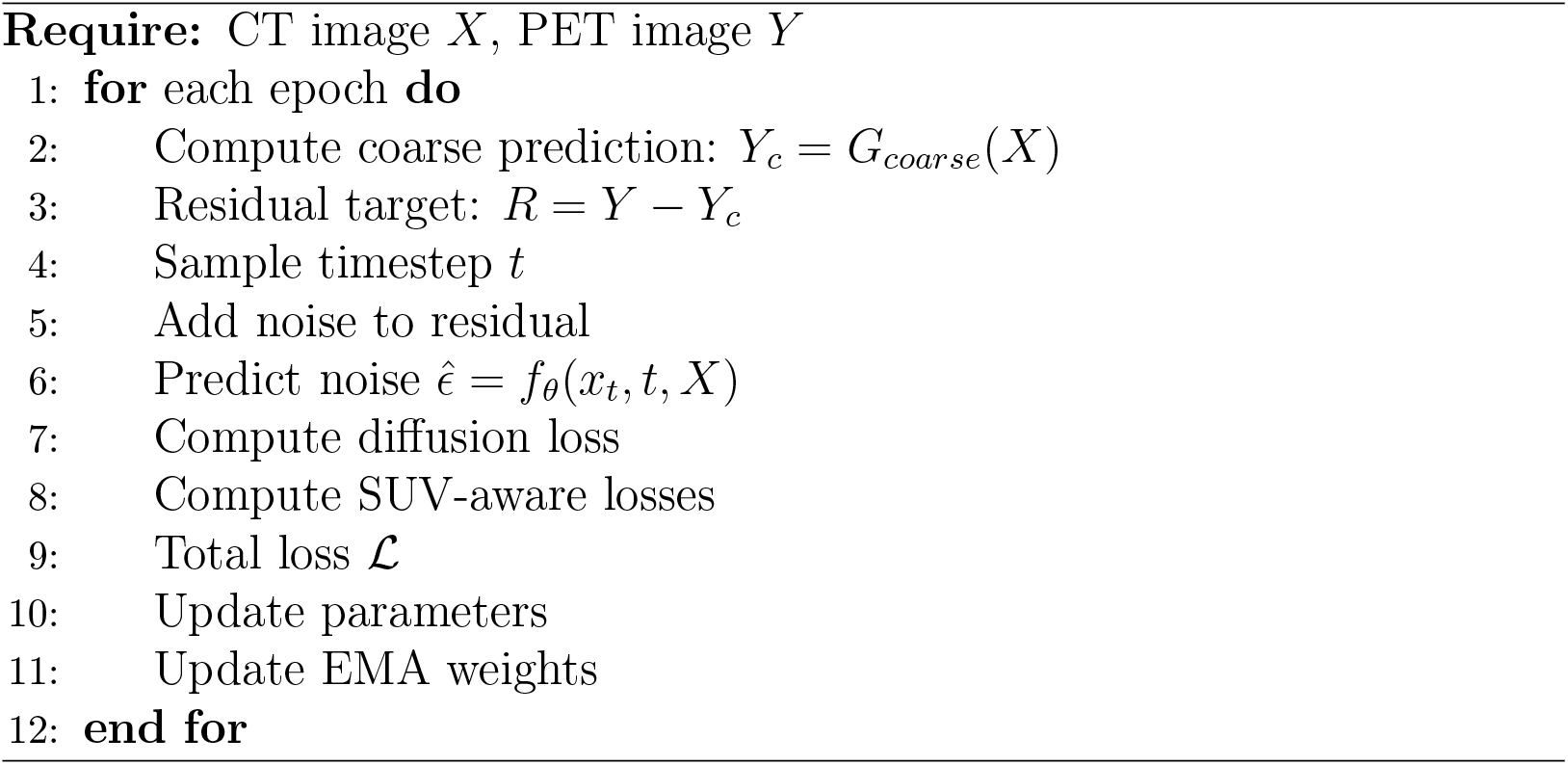

#### Algorithm 2 Inference Algorithm

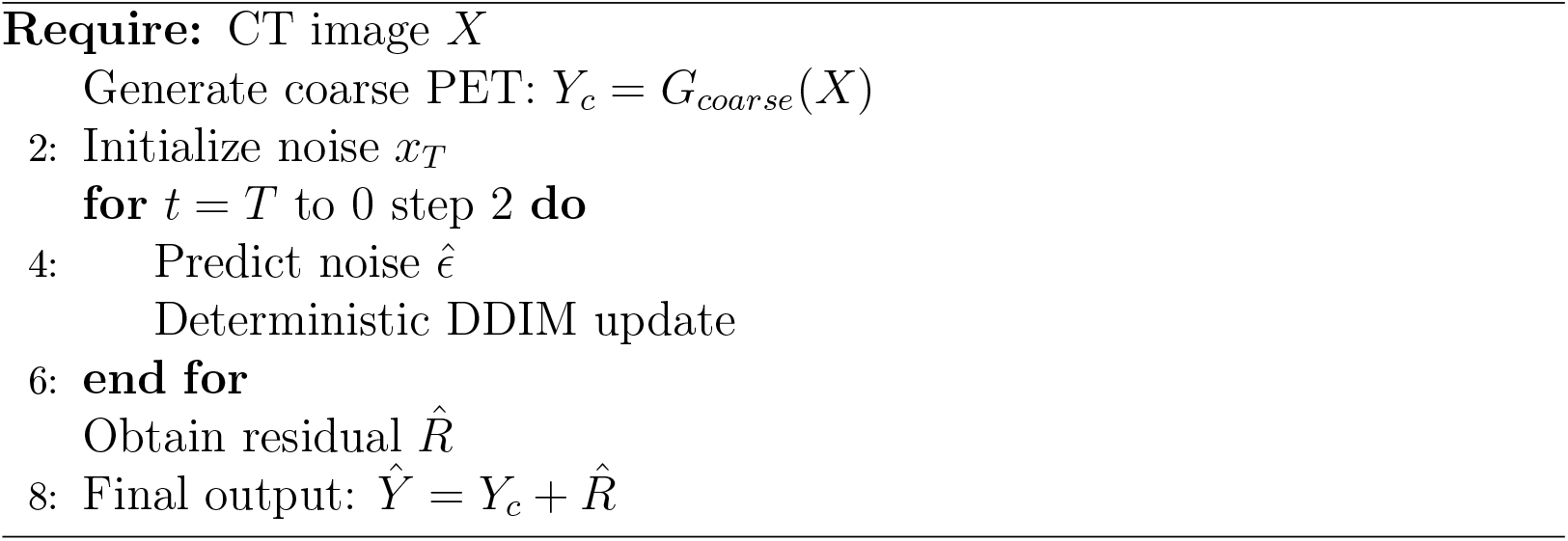

## 4. Experiments

### 4.1. Datasets

The study evaluates the proposed framework on two paired CT–PET datasets to assess both in-domain performance and cross-domain generalization. This work was performed on the publicly available CT–PET paired dataset from Kaggle (Khan, 2023) and on a subset of the CT–PET dataset introduced by Nguyen et al. (Nguyen et al., 2025). The Kaggle CT-PET dataset was split into 3,984 training images, 830 validation images, and 747 testing images. The CPDM subset used in our experiments contains 172 training images, 37 validation images, and 38 testing images.

### 4.2. Evaluation Metrics

To comprehensively evaluate both structural realism and metabolic fidelity, this work reports SSIM, MAE, SUVmean Error, SUVmax Error, TBR Log Error, Leison Dice, and LPIPS. Structural Similarity Index (SSIM) measures structural similarity between predicted and ground-truth PET images, focusing on luminance, contrast, and structural components. Mean Absolute Error (MAE) quantifies pixel-wise intensity differences between synthetic and real PET images. SUVmean Error measures absolute difference in average standardized uptake value (SUV) across the entire scan, reflecting global metabolic consistency while SUVmax Error measures error in maximum SUV value, critical for tumor staging and therapy response evaluation. TBR Log Error evaluates tumor-to-background ratio alignment using logarithmic difference to stabilize scale variations. Lesion Dice measures overlap between predicted and ground-truth lesion masks obtained using SUV thresholding. Learned Perceptual Image Patch Similarity (LPIPS) assesses perceptual similarity using deep feature representations.

## 5. Results

### 5.1. In-Domain Evaluation

Table 2 reports performance when trained and evaluated on the same dataset.

**Table 1:** Dataset Statistics.

| Dataset | Train | Validation | Test |
| --- | --- | --- | --- |
| Kaggle CT-PET (Khan, 2023) | 3984 | 830 | 747 |
| CPDM Subset (Nguyen et al., 2025) | 172 | 37 | 38 |

**Table 2:** In-domain performance of the proposed model.

| Dataset | SSIM | MAE | SUVmax Err | TBR Err | Dice | LPIPS |
| --- | --- | --- | --- | --- | --- | --- |
| Kaggle CT-PET (Test) | 0.8109 | 0.2199 | 1.8180 | 0.4001 | 0.0261 | 0.1974 |
| Kaggle CT-PET (Val) | 0.8426 | 0.1596 | 1.2947 | 0.5261 | 0.0109 | 0.1681 |
| CPDM subset (Test) | 0.7113 | 0.2827 | 0.2391 | 0.8690 | 0.2055 | 0.2758 |
| CPDM subset (Val) | 0.7132 | 0.2728 | 0.2211 | 0.8452 | 0.2196 | 0.2683 |

**Table 3:** Cross-domain evaluation results.

| Train → Test | SSIM | MAE | SUVmax Err | Dice |
| --- | --- | --- | --- | --- |
| Kaggle CT-PET → CPDM subset | 0.7214 | 0.2801 | 2.4675 | 0.1477 |
| CPDM subset → Kaggle CT-PET | 0.7968 | 0.2248 | 2.8602 | 0.0230 |

**Table 4:**
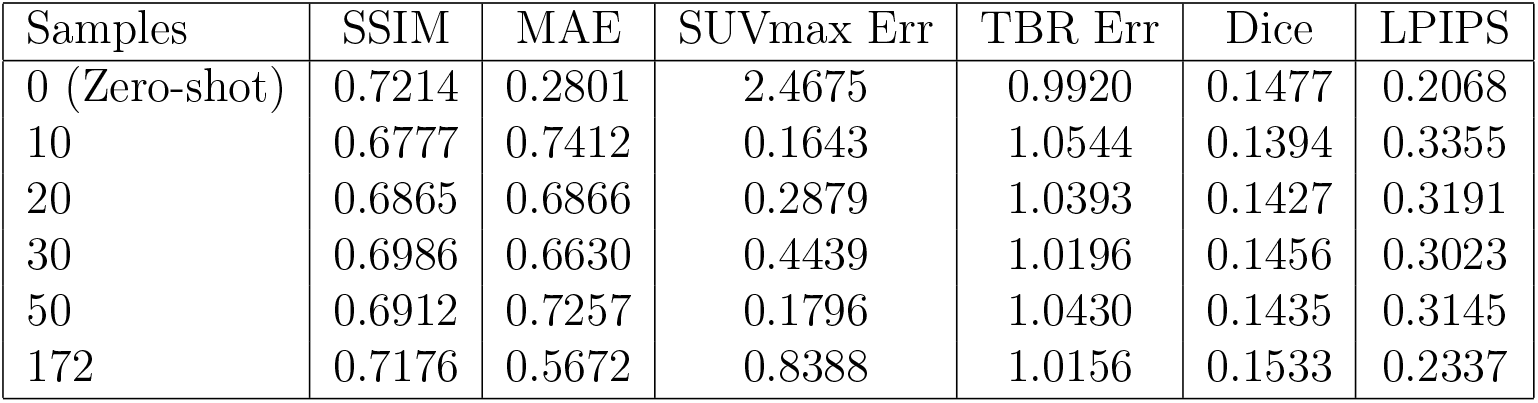
Few-shot adaptation results on CPDM test set.

| Samples | SSIM | MAE | SUVmax Err | TBR Err | Dice | LPIPS |
| --- | --- | --- | --- | --- | --- | --- |
| 0 (Zero-shot) | 0.7214 | 0.2801 | 2.4675 | 0.9920 | 0.1477 | 0.2068 |
| 10 | 0.6777 | 0.7412 | 0.1643 | 1.0544 | 0.1394 | 0.3355 |
| 20 | 0.6865 | 0.6866 | 0.2879 | 1.0393 | 0.1427 | 0.3191 |
| 30 | 0.6986 | 0.6630 | 0.4439 | 1.0196 | 0.1456 | 0.3023 |
| 50 | 0.6912 | 0.7257 | 0.1796 | 1.0430 | 0.1435 | 0.3145 |
| 172 | 0.7176 | 0.5672 | 0.8388 | 1.0156 | 0.1533 | 0.2337 |

### 5.2. Cross-Domain Generalization

To evaluate robustness across imaging domains, models trained on one dataset were tested on the other.

Cross-domain transfer significantly increases SUVmax error, indicating strong domain sensitivity in metabolic intensity calibration.

### 5.3. Few-Shot Rural Adaptation

To simulate realistic rural deployment, the pretrained model of Kaggle CT-PET dataset was finetuned using progressively increasing subsets of CPDM training data (10, 20, 30, 50, and 172 samples). All models were evaluated on the held-out CPDM test set.

### 5.4. Qualitative Comparison of Synthetic PET Reconstruction

Figure 2 presents a qualitative comparison between the input CT image (HU-normalized), the ground-truth PET image in SUV space, the in-domain predicted PET (fine-tuned model), and the zero-shot predicted PET (cross-domain inference). The CT image (left panel) provides anatomical structural information without metabolic activity. The ground-truth PET image (second panel) demonstrates focal regions of elevated glucose metabolism, highlighted within the cyan bounding box. These high-intensity regions correspond to clinically relevant metabolic hotspots. The in-domain predicted PET (third panel), obtained after fine-tuning on the CPDM dataset, successfully reconstructs the spatial distribution and intensity profile of the focal lesion. The lesion morphology and contrast relative to surrounding tissue are preserved, indicating effective adaptation to domain-specific SUV characteristics.

**Figure 1.**
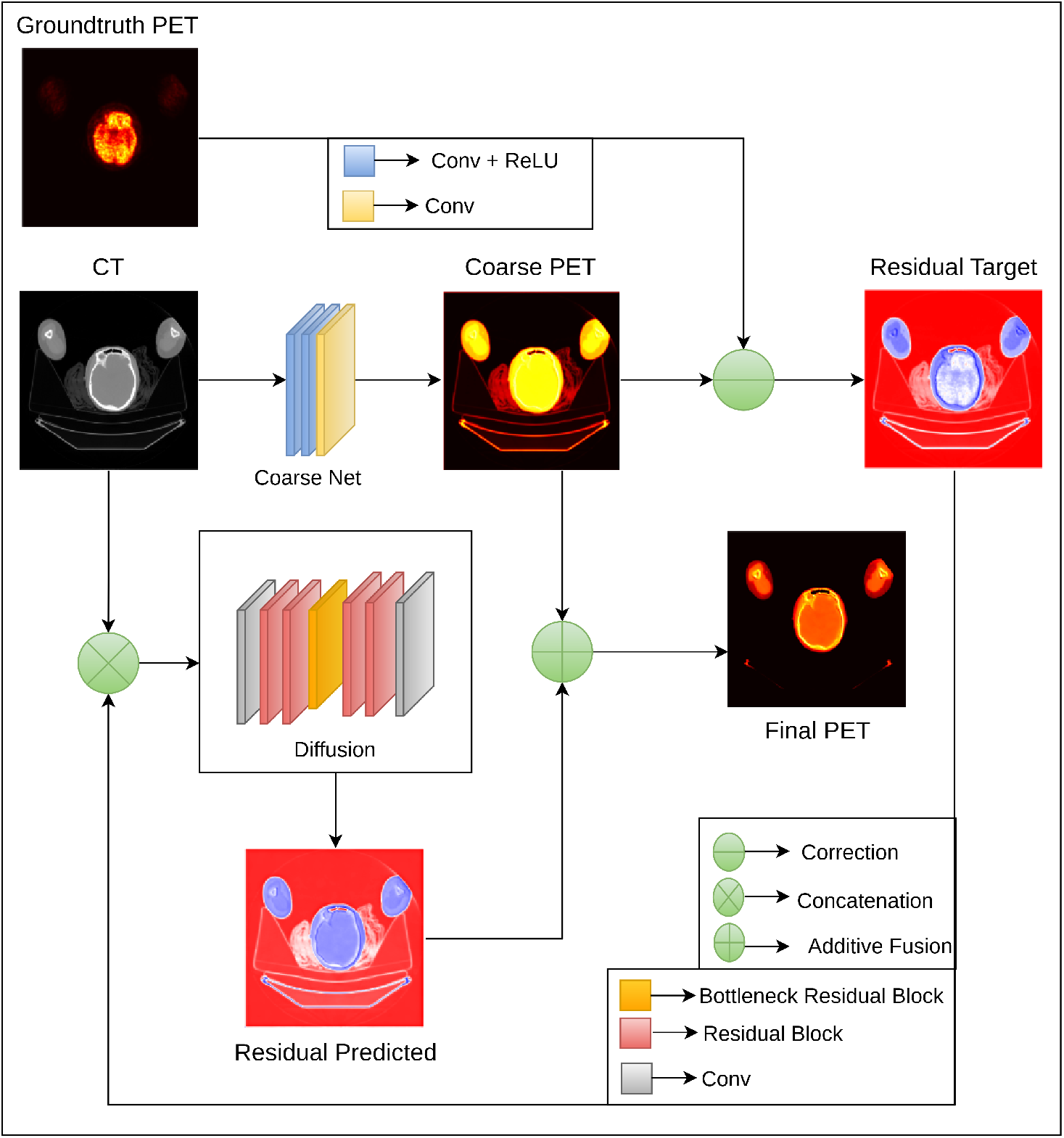
Conditional Diffusion Framework from CT to PET translation.

**Figure 2.**
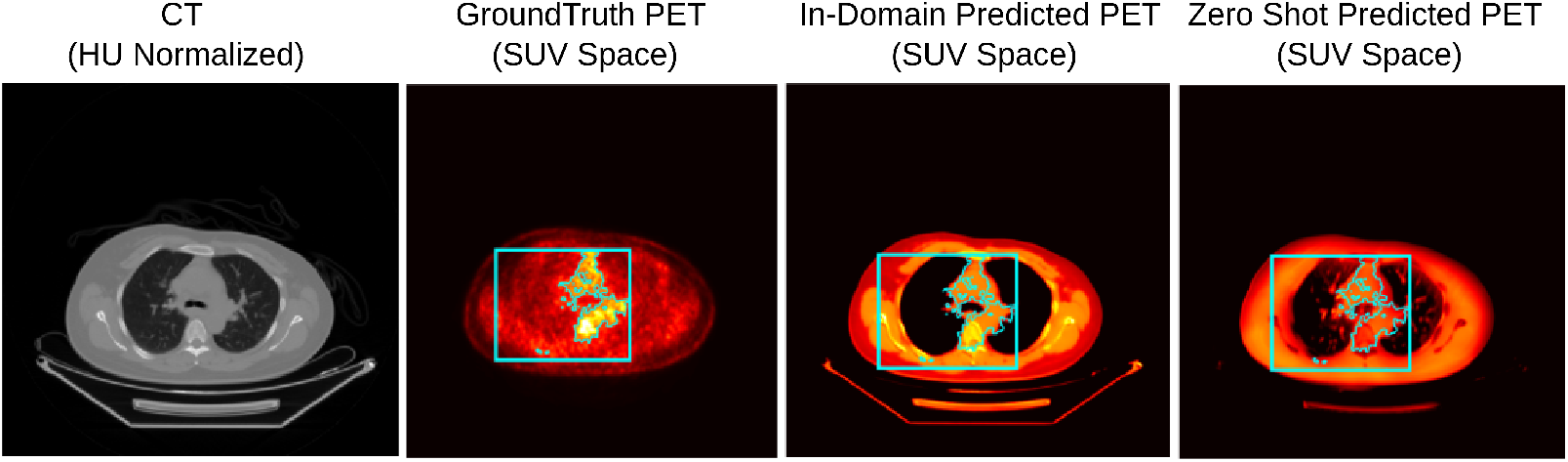
Qualitative comparison of CT input, ground-truth PET, in-domain predicted PET, and zero-shot predicted PET in SUV space. Cyan bounding box highlights focal metabolic region.

In contrast, the zero-shot predicted PET (fourth panel), generated without domain-specific fine-tuning, captures the general anatomical-metabolic correspondence but underestimates lesion contrast and exhibits reduced intensity calibration. While structural localization remains plausible, the SUV distribution appears compressed compared to the in-domain prediction. Importantly, both synthetic outputs preserve the overall lesion location, but fine-tuning significantly improves metabolic contrast fidelity. This qualitative evidence supports the hypothesis that domain adaptation enhances quantitative SUV consistency and improves downstream referral simulation reliability in rural deployment scenarios.

#### Algorithm 3 Training Algorithm

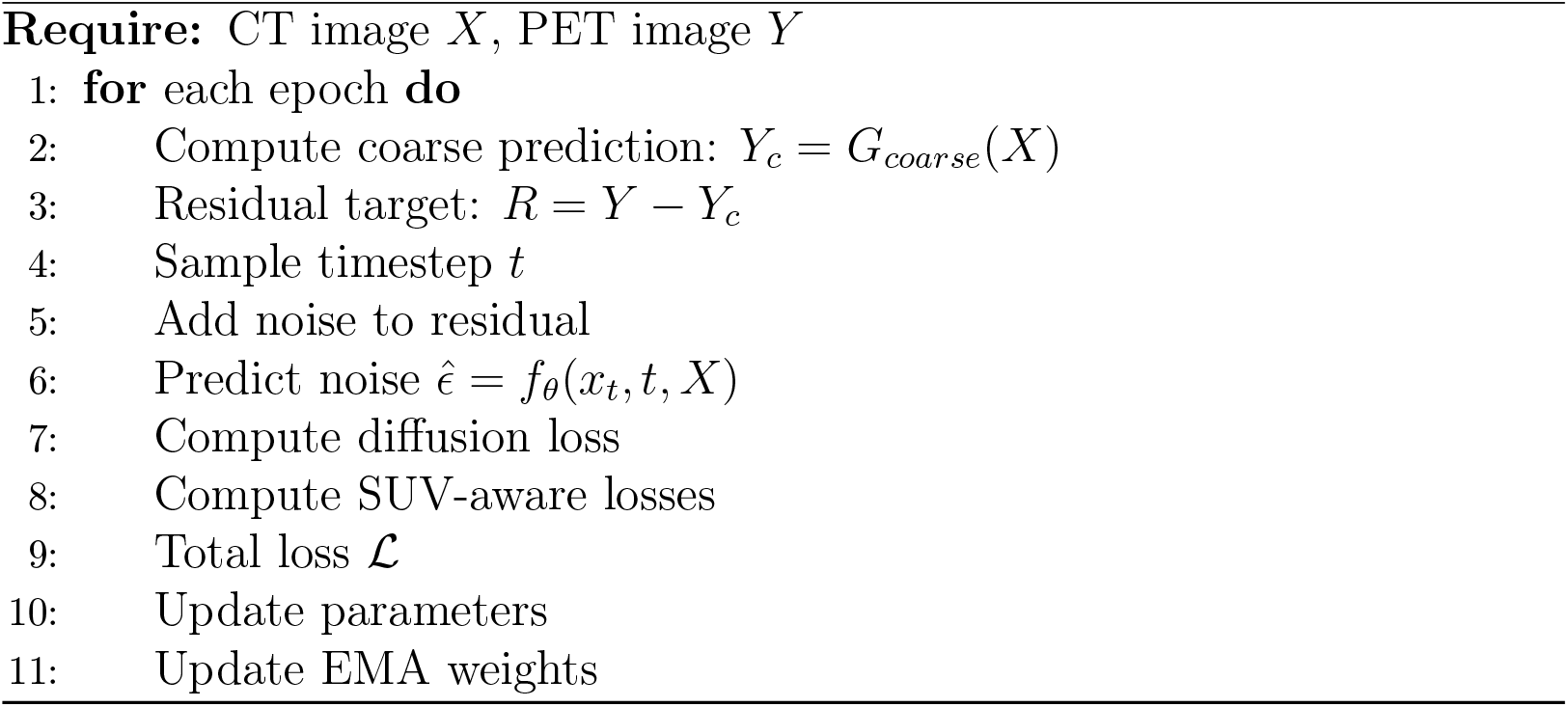

#### Algorithm 4 Inference Algorithm

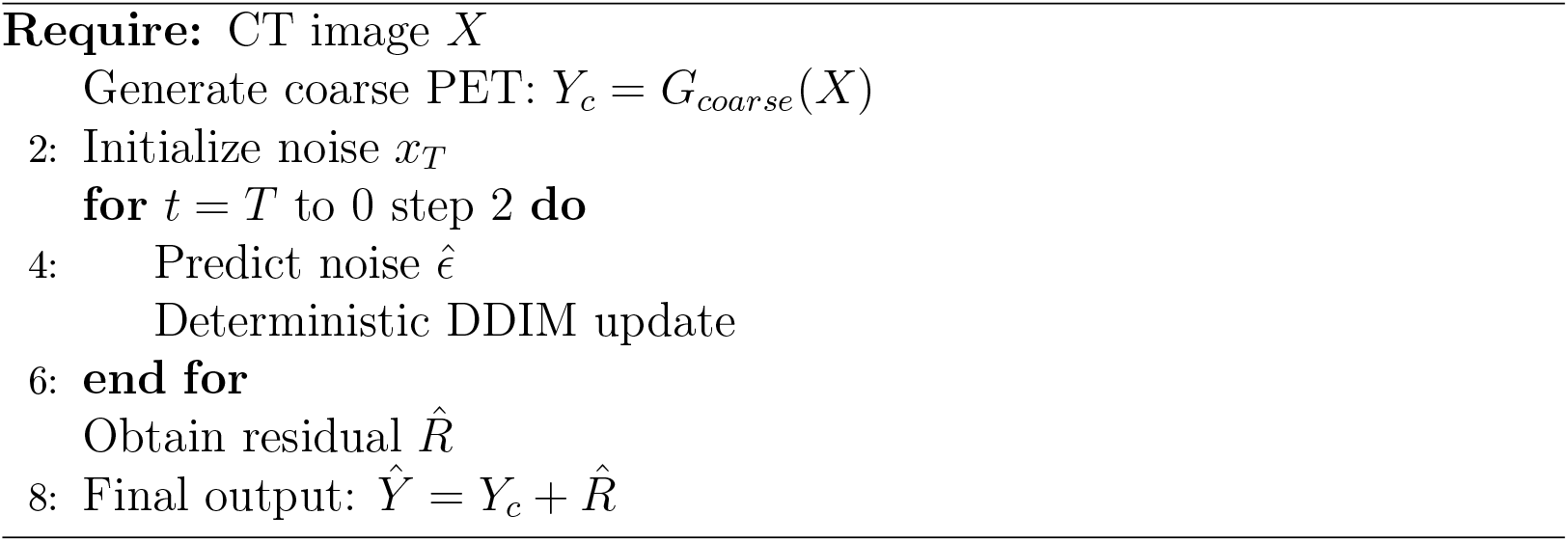

## 6. Discussion

### 6.1. Domain Sensitivity and Calibration

Zero-shot transfer from Kaggle CT-PET to CPDM subset resulted in substantial SUVmax underestimation (2.47 error), highlighting the domain sensitivity of PET intensity distributions. Differences in scanner calibration, reconstruction algorithms, and radiotracer protocols contribute to this discrepancy.

### 6.2. Few-Shot Metabolic Recalibration

Few-shot fine-tuning dramatically reduced SUVmax error, with as few as 10 local samples decreasing error below 0.2. This indicates that limited local data is sufficient to recalibrate metabolic intensity distributions. While global MAE increased due to intensity redistribution, peak uptake preservation improved substantially.

### 6.3. Implications for Rural Healthcare

In rural healthcare settings where PET scanners are limited, the goal is not pixel-perfect reconstruction but metabolic risk stratification. The results demonstrate that:

- Zero-shot deployment is insufficient due to domain mismatch.
- Few-shot adaptation rapidly corrects SUV scaling.
- Limited local datasets (10–50 cases) are adequate for practical calibration.

This supports a realistic deployment strategy involving pretraining on comparatively large public datasets followed by lightweight local fine-tuning.

## 7. Rural Healthcare Deployment Strategy

In this work, the proposed CT-to-PET translation framework is positioned not as a replacement for clinical PET imaging, but as a metabolic screening and referral support tool. Based on experimental findings, this work proposes the following practical rural deployment strategy:

1. Large-Scale Pretraining: Train the diffusion-based model on publicly available datasets to learn general anatomical-to-metabolic priors.
2. Local Calibration (Few-Shot Fine-Tuning): Collect a small number (10–50) of locally acquired paired CT–PET scans and fine-tune the pretrained model to adjust SUV intensity scaling and scanner-specific characteristics.
3. Screening Deployment: Use the adapted model to generate synthetic PET from routine CT scans for:
  - Metabolic risk stratification,
  - SUV-based referral prioritization,
  - Preliminary tumor burden assessment.

### 7.1. Operational Workflow in Rural Settings

For each incoming CT scan:

1. Generate synthetic PET using the fine-tuned model.
2. Compute SUV-based indicators (e.g., SUV_*max*_, tumor-to-background ratio).
3. Apply predefined referral thresholds.
4. Refer high-risk cases to centralized PET facilities.

This approach enables resource-aware triage while preserving specialist diagnostic authority.

### 7.2. Key Experimental Evidence

Few-shot experiments demonstrate that even limited local data (10–30 cases) substantially reduces SUV_*max*_ error compared to zero-shot transfer. This confirms that domain-specific intensity calibration can be achieved with minimal additional data, making deployment feasible in low-resource environments. Importantly, the framework emphasizes peak uptake preservation, which is clinically relevant for oncology referral decisions.

To evaluate real-world deployment feasibility, the study simulates a rural referral workflow using SUV-based decision rules on synthetic PET outputs. For a representative CPDM subset test slice (ID 0016) after 50-sample fine-tuning, the model produced:

- Predicted SUV_*max*_ = 5.67
- Tumor-to-Background Ratio (TBR) = 5.29
- Lesion Fraction = 18.4%
- Final Decision: REFER TO PET CENTER

Although SUV_*max*_ did not exceed the absolute referral threshold of 6.0, the high TBR and substantial focal metabolic region triggered a referral decision under the screening policy. This demonstrates that the proposed framework does not rely solely on peak intensity, but preserves clinically meaningful metabolic contrast patterns that can guide triage decisions. Importantly, such decisions can be made using only CT-derived synthetic PET, potentially enabling early identification of high-risk patients in rural settings without immediate access to PET scanners.

## 8. Conclusion

This work proposed a rural-optimized conditional diffusion framework for CT-to-PET translation that integrates residual learning and SUV-aware optimization. The model demonstrates strong in-domain metabolic consistency and reveals domain sensitivity under zero-shot transfer. Few-shot adaptation experiments show that limited local data (10-50 paired cases) is sufficient to substantially reduce SUV_*max*_ error, enabling practical metabolic recalibration. These findings support a realistic deployment strategy involving large-scale pretraining followed by lightweight local fine-tuning. Rather than replacing PET imaging, the proposed framework serves as a screening and referral support tool in PET-limited rural settings. By bridging structural CT availability with approximate metabolic assessment, this approach contributes toward improving equitable access to oncology imaging support in resource-constrained environments.

### Limitations

Cross-domain zero-shot performance reveals significant sensitivity to SUV calibration differences across datasets. This highlights the necessity of local fine-tuning prior to deployment. Although SUV_*max*_ preservation improves with few-shot adaptation, global MAE and perceptual similarity metrics may degrade due to intensity redistribution. This reflects a trade-off between metabolic calibration and pixel-level reconstruction accuracy. Lesion Dice scores remain moderate, as tumor segmentation is indirectly derived from SUV thresholding rather than expert-annotated masks. Future work should incorporate lesion-aware supervision and multi-task optimization. Finally, this framework is not intended to replace clinical PET imaging. Synthetic PET outputs should be interpreted as screening-level estimations rather than definitive diagnostic measurements.

## Data Availability

The Kaggle CT-PET dataset is taken from \cite{kaggle_ctpet_dataset} and the CPDM subset is taken from \cite{nguyen2025ct}.} [mentioned in the manuscript]

## Acknowledgment

This work received no external funding.

